# Impact of an environmental epidemiology board game on knowledge, experience, and attitudes among children: A pilot study

**DOI:** 10.1101/2021.05.13.21256979

**Authors:** Shelly Melissa Pranic, Tina Batinovic

## Abstract

Children are especially vulnerable to environmental harms, thus increasing their knowledge about risk factors caused by environmental exposures, in addition to epidemiology by providing practical experiences through a non-computer based educational game in environmental epidemiology may lead to more promising behavioral outcomes. Because no educational game exists in environmental epidemiology, we augmented an existing epidemiology game by adding an environmental health theme, involving practical decision-making and problem solving in environmental epidemiology. A cross-sectional survey assessed the game’s impact, on children’s knowledge, experience, and attitudes regarding environmental health, and experience and attitudes toward epidemiology following game exposure in 2016. Participants from Croatia had a median age of 11.5 years with interquartile range (IQR) 11-13. The majority of children indicated they had learned a lot about the environmental health concepts newly added to an existing epidemiology game. Our modified game offers interactive and practical examples, which may facilitate teaching environmental epidemiology to elementary school students.

## Introduction

In the context of increased environmental stressors affecting children, education about environmental health and awareness of environmental factors that can adversely affect their health is important. As children are especially vulnerable to environmental detriments (Quiros-Alcala et al. 2011; Tuntawiroon et al. 2007; Wipfli et al. 2008), environmental health education offers children basic information needed to gather knowledge about the environment as well as to mitigate exposure to environmental risk factors (Boeve-de Pauw et al. 2015; Claudio et al. 1998). Environmental health education for elementary and secondary school children is effective in that children’s knowledge about the environment from the classroom translates into increased awareness about the environment (Quiros-Alcala et al. 2011). Recent studies showed the inclusion of content-specific as well as pedagogical approaches for the inclusion of environmental sustainable development in the curriculum of elementary school children improved the children’s realistic application of their knowledge and skills (Boeve-de Pauw et al. 2015; Cincera and Maskova 2011). Moreover, children’s attitudes, behaviors, and misconceptions that are acquired during early childhood may be determinate, thus before children grow older, it would be beneficial to introduce activities that foster critical thinking (Duschi et al. 2007), such as a serious game.

Inciting positive behaviors toward the environment is not an easy task as exclusive barriers or other factors may influence how knowledge about the environment translates into practice (Heimlich and Ardoin 2008; Kollmuss and Agyeman 2002). Studies have found many complex intermediators between knowledge or attitude and behavior, which modulate how individuals apply learned concepts (Mayer 2019; Saphores et al. 2012). Further, knowledge of problems and solving them concerning the environment were associated with positive behaviors toward the environment (Bamberg and Möser 2007; Hines et al. 1987). Educational games or “serious games” that target adults, children, and adolescents convey information to initiate behavioral change and exist for population-specific problems in diverse fields such as education, business, marketing, healthcare, and environmental management (Drummond et al. 2017; Gilliam et al. 2016; Sawyer 2007). Educational games played in the classroom provide a promising way to deliver behavior-modifying education depending on the content and design of the game (Gilliam et al. 2016; Viggiano et al. 2018). There are two major types of mediums used for serious games to convey information: computer- or non-computer based (Lean et al. 2006). Computer-based serious games prompt users to follow game objectives with computer simulations whereas non-computer or table based serious games present objectives in paper form (Lean et al. 2006).

Most of the literature about serious games predominantly concerns computer- or digital-based games or games in digital form; nevertheless, serious games regardless of their form encompass real-world problem solving for fictitious scenarios where players as decision-makers strive toward achieving the goals of the game (Annetta 2010; Squire 2008). A recent systematic review highlighted the effectiveness of digital serious games on decision-making and planning surrounding environmental issues to modulate behavior after learning (Morganti et al. 2017). According to this systematic review, serious games encourage children as well as adult game players toward pro-environmental behaviors after playing the game (Morganti et al. 2017). Regarding board games, a recent study showed that children highly accepted a classroom board game intended for sexuality education and that introduction of the game into the curriculum was feasible (Gilliam et al. 2016). A recent pilot study of a board game for adolescents increased their awareness about tobacco companies’ marketing approaches and harms from smoking (Gilliam et al. 2019). Serious games have the ability to turn children’s knowledge into behavioral change (Madani et al. 2017). Currently, there are no evaluations of the knowledge and perceptions of elementary school students about educational non-computer based serious games in environmental epidemiology with a pedagogical foundation.

Accordingly, we modified the “What’s Lurking in Lunch?” board game by Arizona State University educators originally created to teach epidemiology to elementary school children (Science Center 2014). Our modified game included scenarios in which the children role-played as environmental epidemiologists tracking the source of an environmental contaminant when presented with a patient exhibiting a specific set of symptoms (Supplementary Table 1). Our game includes the epidemiological learning objectives of the original game in terms of observation of the problem, information synthesis, and interpretation of findings (Science Center 2014). However, we modified the game to include environmental health topics in addition to the original epidemiology-based game theme. In line with our modifications, we named our game “What’s Lurking in that Building?”

Studies thus far have focused on the general inclusion of environmental educational material in schools’ curricula and recently on the development of an interactive digital game in which users of various ages increased their learning and interest in environmental education (Su 2018). We propose that a non-digital game with an environmental epidemiology basis would encourage students to learn more about and solve problems concerning environmental epidemiology. Specifically, we aimed to determine the 1) experiences and attitudes of students regarding environmental health and epidemiology as subjects in school and 2) knowledge and attitudes surrounding the non-digital “What’s Lurking in that Building?” game.

## Methods

The cross-sectional pilot study took place during a workshop at the University of Split’s Festival of Science on April 21, 2016. The 2-hour workshop conducted in English consisted of: 1) a 10-minute talk about environmental health; 2) a modified game to introduce students to environmental epidemiology; and 3) the construction of a green building. First, one author (S.P.) gave a 10-minute talk about environmental health in terms of detriments and sustainability based on the freely-available materials on the Environmental Health Student Portal from the U.S. National Library of Medicine (U.S. National Library of Medicine 2016) along with examples of common indoor (National Institute of Environmental Health Sciences 2016a) and outdoor irritants (National Institute of Environmental Health Sciences 2016b) from the U.S. National Institute of Environmental Health Sciences. The talk specified the importance of conserving water, electricity, and waste recycling according to recommendations for children from the U.S. National Institute of Environmental Health Sciences (National Institute of Environmental Health Sciences 2011).

Children played our modified “What’s Lurking in Lunch?” food borne illness game for our pilot study [see Supplementary Table 1] (Science Center 2014). The learning objectives of the “What’s Lurking in that Building?” game allowed the children to: 1) compose relevant questions through their own personal experiences with environmental contaminants; 2) conceive an investigation into the causes of illness based on their composed questions; and 3) connect exposures to certain environmental contaminants to health outcomes. The players needed to discover 1) the causes of the illness from improper handling of environmental contaminants; 2) the organism or substance that caused the illness; 3) the origin of the infection; 4) contaminant causing an outbreak; and 5) how to calculate the overall attack rate (number of new individuals affected by an illness divided by the total exposed population) (Science Center 2014).

Finally, the students constructed their own environmentally friendly green buildings using stationery materials during the workshop. Students were encouraged to incorporate energy conserving elements into their buildings, as learned from the talk about environmental sustainability. After the board game and model green building construction, all (n=13) of the workshop participants completed a survey (see Supplementary Material, Questionnaire) to assess their experiences and attitudes regarding environmental health or epidemiology courses in their elementary school curricula.

### Instrument

We collected responses on participants’ experiences and attitudes regarding courses in environmental health and epidemiology from participants with a 9-item survey (see Supplementary Material, Questionnaire). We modified the survey created by Gilliam and colleagues (Gilliam et al. 2016) with pertinent questions for use in the current study. The survey for the current study gathered data on the students’ experiences and attitudes surrounding environmental health and epidemiology, their opinion of and knowledge gained from the “What’s lurking in that Building?” game (see Supplementary Material, Questionnaire).

The original English language survey was translated into Croatian by the second author (T.B.). In order to ensure accurate translation, the Croatian version was subsequently back-translated into English by a bilingual research scientist (Brislin 1970). The authors compared the original English and the back-translated English versions of the survey for translation accuracy. We did not translate any parts of the “What’s Lurking in that Building?” game into the Croatian language; however, the reading level of the players’ materials was least at a 3^rd^ grade English reading level (Science Center 2014). Therefore, the fifth to eighth grade students who participated in the workshop were able to read the content of the game, as English reading and writing courses are mandatory in the Croatian elementary school curriculum from the first to the eighth grade (Croatian Ministry of Science, Education and Sports 2006).

We followed the Strengthening the Reporting of Observational Studies in Epidemiology (STROBE) guidelines (von Elm et al. 2007) and obtained approval from the Ethics Committee at the University of Split School of Medicine for this study.

### Statistical Analysis

We performed all analyses using MedCalc Statistical Software version 17.1 (MedCalc Software bvba, Ostend, Belgium). We reported descriptive statistics for demographic data, environmental health and epidemiology experiences and attitudes, and knowledge of and attitudes about the game as percentages or medians (interquartile range [IQR]) with corresponding 95% confidence intervals (CIs).

## Results

### Participant description

Thirteen children participated in the study from three elementary schools, with 11 (84.6%) from Split 3 Elementary School and one (7.7%) each from Mertojak and Pojisan schools from the community of Split, Croatia. Their median age was 11.5 years (IQR 11-13). For the 2015/16 school year, the median grade was 6 (IQR 5-7). Mostly, children were in the sixth grade (n = 5, 30.8%), followed by four (38.5%) in the fifth grade, 3 (23.1%) in the seventh grade, and one (7.7%) in the eighth grade.

### Experience with environmental health or epidemiology

Most of the children reported having prior experience with environmental health as they reported having environmental health separately from a biology course with a median rating of 4 (IQR 4-4, 95% CI 3.7-4.0) shown in Table 1. Notably, all 11 (84.6%) of the children that reported learning about environmental health aside from biology were from Split 3 Elementary School (Table 1). Two (15.4%) students reported that they never had a course in school regarding environmental health (one from Pojisan and another from Mertojak Elementary Schools) shown in Table 1.

**Table 1.**
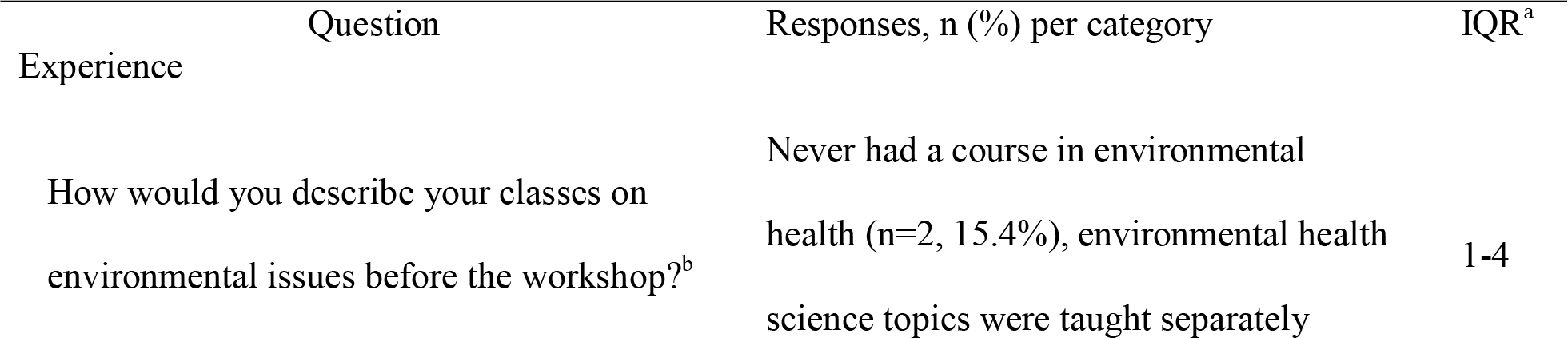
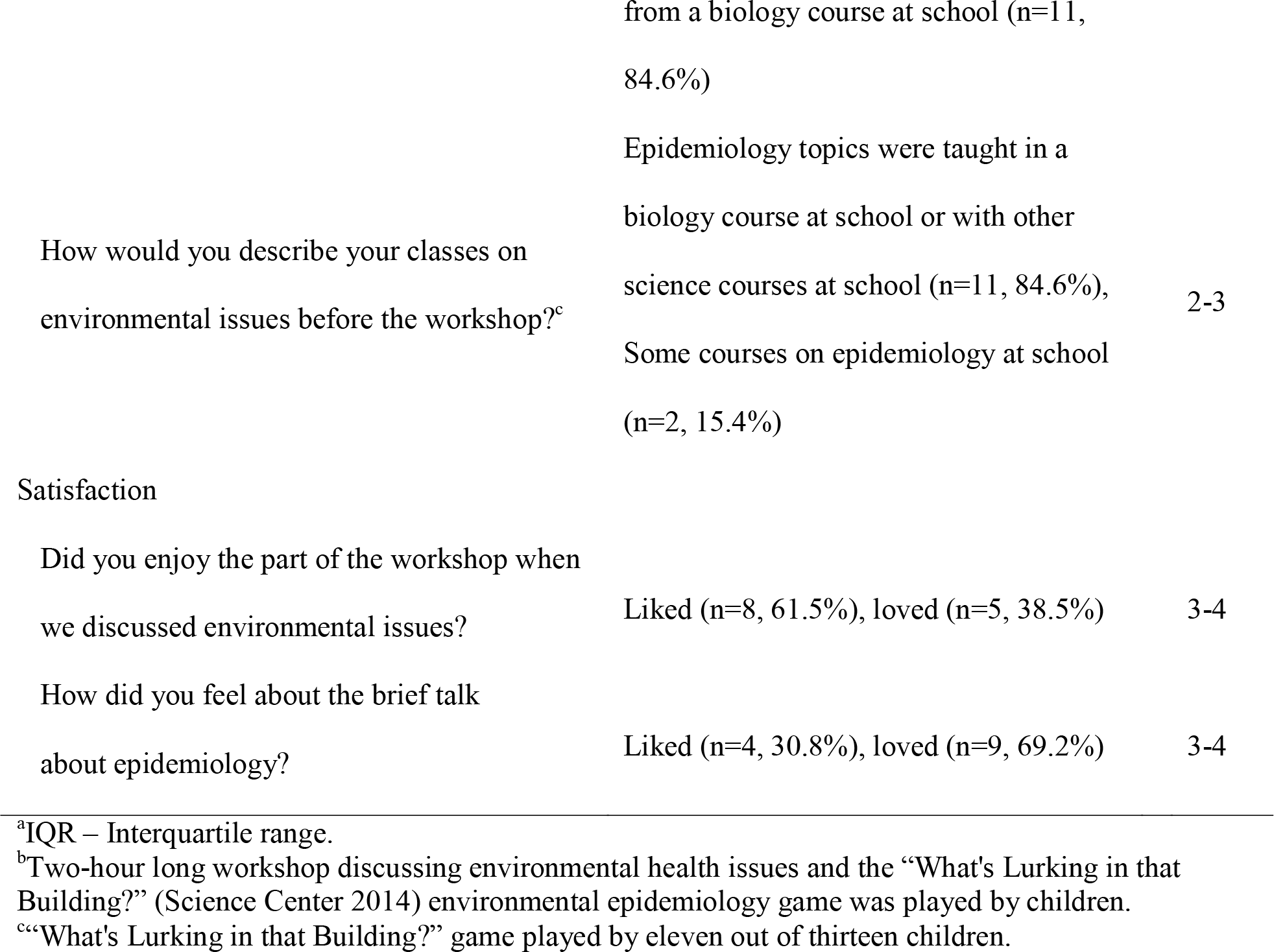
Elementary school students’ experience and satisfaction with environmental health or environmental issues after a workshop and the “What’s Lurking in that Building?” game on environmental epidemiology.

| Experience | Question | Responses, n (%) per category | IQR<sup>a</sup> |
| --- | --- | --- | --- |
|  | How would you describe your classes on environmental issues before the workshop?<sup>b</sup> | Never had a course in environmental health (n=2, 15.4%), environmental health science topics were taught separately | 1-4 |

|  |  |  |
| --- | --- | --- |
|  | from a biology course at school (n=11, 84.6%) |  |
| How would you describe your classes on environmental issues before the workshop?<sup>c</sup> | Epidemiology topics were taught in a biology course at school or with other science courses at school (n=11, 84.6%),<br>Some courses on epidemiology at school (n=2, 15.4%) | 2-3 |
| Satisfaction |  |  |
| Did you enjoy the part of the workshop when we discussed environmental issues? | Liked (n=8, 61.5%), loved (n=5, 38.5%) | 3-4 |
| How did you feel about the brief talk about epidemiology? | Liked (n=4, 30.8%), loved (n=9, 69.2%) | 3-4 |

<sup>a</sup>IQR – Interquartile range.
<sup>b</sup>Two-hour long workshop discussing environmental health issues and the “What's Lurking in that Building?” (Science Center 2014) environmental epidemiology game was played by children.
<sup>c</sup>“What's Lurking in that Building?” game played by eleven out of thirteen children.

Predominantly, children from Split 3 elementary school learned about epidemiology along with another subject, such as biology with a median rating of 3 (IQR 3-3, 95%CI 2.8-3.0) shown in Table 1. Two students from Pojisan and Mertojak Elementary Schools reported having some prior knowledge about epidemiology from a course at school (Table 1).

### Attitudes about environmental health during the workshop

As shown in Table 1, most children reported that they learned “a lot” from the brief talk in the workshop concerning environmental health. Two students from Split 3 Elementary School reported that they had learned “some useful things” about environmental health from the workshop. Out of the 11 students from Split 3 Elementary School who participated in the “What’s Lurking in that Building?” game, the majority (n = 7, 53.8%) reported that they learned “a lot” while 4 students reported learning “some useful things” (Table 1). All of the 13 children participated in the constructing of a green building exercise and reported that they had learned a lot about environmental health (Table 1). The survey did not collect responses about learning epidemiolo gy as the game already had an epidemiology focus and we wanted to collect information on learning and attitudes concerning the new environmental health content.

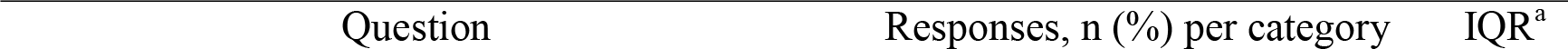

### Attitudes about environmental health and epidemiology

As shown in Table 1, most children liked the environmental health theme of the workshop, with an overall median score of 3 (IQR 3-4, 95% CI 3-4) for this item. Regarding the children’s satisfaction with the epidemiology theme of the workshop, the majority loved the theme with a median rating of 4 (IQR 3-4, 95% CI 3-4) [Table 1].

### Learning from the “What’s Lurking in that Building?” game

Out of the 11 (84.6%) children who provided responses regarding their opinion of the “What’s Lurking in that Building?” game, most reported that they learned a lot with a median rating of 4 (IQR 3-4, 95% CI 3.0-4.0), as shown in Table 1. The two students from Pojisan and Mertojak Elementary Schools did not participate in this portion of the workshop.

Table 2 shows the children’s comments about the brief talk on environmental health and the workshop.

**Table 2.**
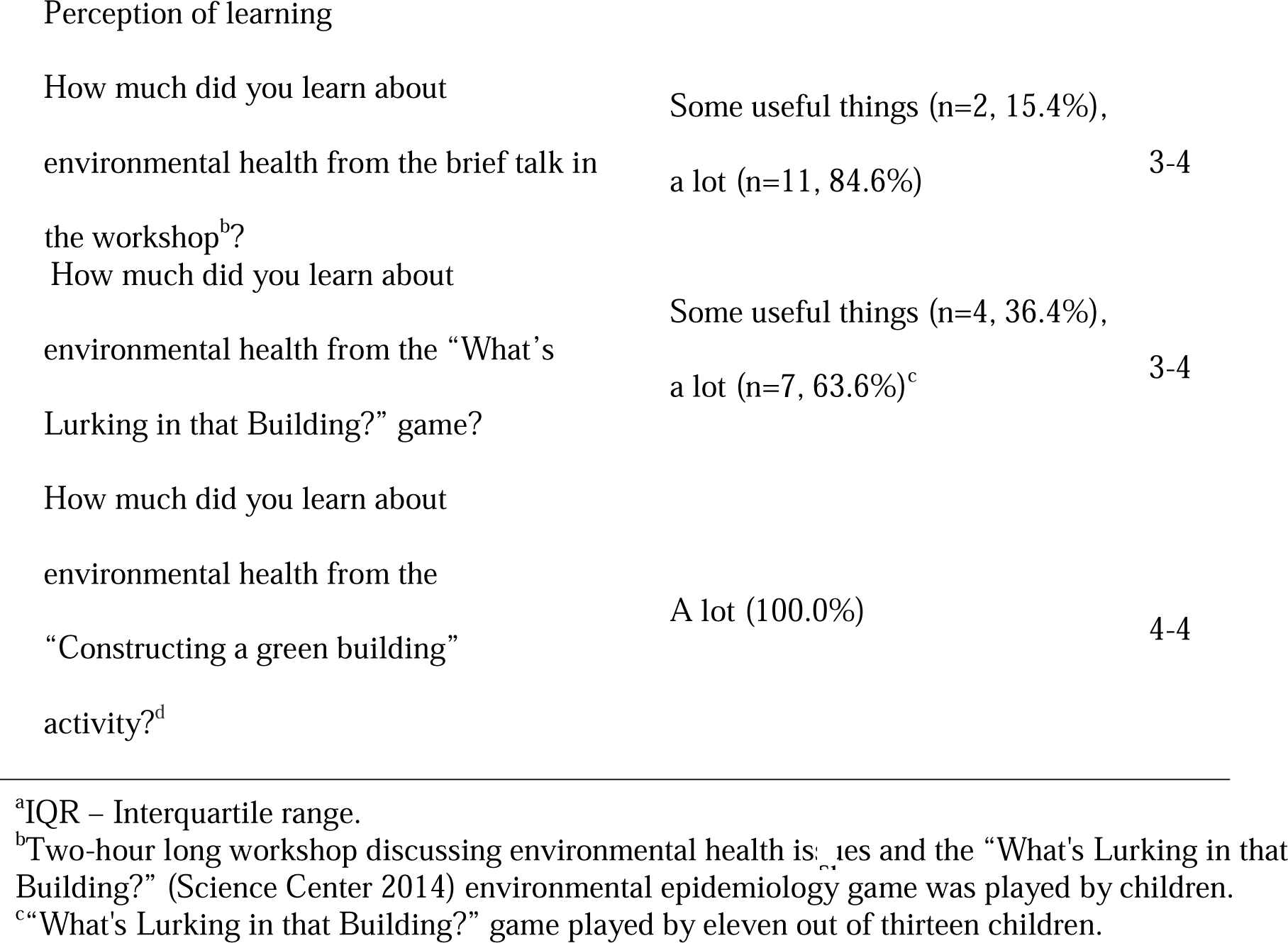
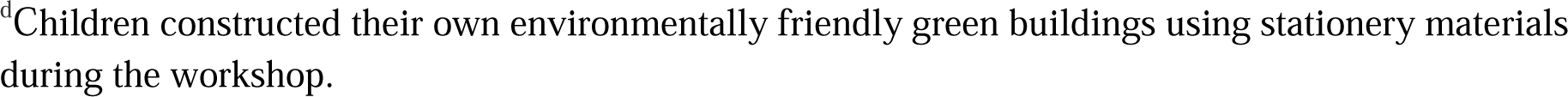
Elementary school students’ perception of learning about environmental health after a workshop and the “What’s Lurking in that Building?” game on environmental epidemiology.

## Discussion

Overall, the elementary school students in our cross-sectional pilot study had experience with environmental health and epidemiology as taught as separate topics apart from biology. The children enjoyed taking part in the workshop that centered on environmental health as well as epidemiology. The children reported that they learned a lot from both the workshop and the “What’s Lurking in that Building?” game that merged the two fields into environmental epidemiology. Based on open-ended commentary, the students enjoyed the practical examples as well as the interactive nature of the workshop in general, with a positive comment about working together during the game. Accordingly, due the enjoyable, interactive nature and real-life practical examples employed by the “What’s Lurking in that Building?” game, the game may help to facilitate in teaching environmental epidemiology to elementary school students.

Evidence from observational and experimental studies in various scientific disciplines showed that when a serious game employs content with both entertainment and educational value, learning is more effective because students are engaged and motivated to learn (Blakely et al. 2009; Cheng and Annetta 2012; Knight et al. 2010; Sitzmann 2011). Currently, there are no studies on the effectiveness of non-computer game-based curricula in the field of environmental epidemiology. In regards to environmental education, Su created an interactive digital serious game that guided players with an entertaining and user-friendly digital interface to make decisions and solve problems concerning the environment (Su 2018). However, Su’s computer game included learning and subsequent application in solely environmental education to adolescents as well as adults, while the non-computer based game used in this study would serve as a novel approach in teaching environmental epidemiology to elementary school children, especially in settings where creating digital games may be unfeasible. In the absence of serious games in environmental epidemiology, a serious game promises to increase knowledge about environmental epidemiology as classroom-based interventions have translated into increased knowledge in environmental education (Phan Hoang and Kato 2016; Vaughan et al. 2003). Phan Hoang and colleagues increased elementary schoolchildren’s knowledge about plastic waste management after an environmental education workshop (Phan Hoang and Kato 2016). Environmental education provided by a trained teacher to Costa Rican schoolchildren resulted in higher environmental literacy in students that transferred to parents and members of the community (Vaughan et al. 2003).

In the future, we imagine that a game that encompasses content specific and pedagogical approaches to teaching environmental epidemiology to students would be advantageous for both schools and students. School administrators would provide a method for their students to become more environmentally conscious in their school-aged years and beyond. Children would learn more about environmental detriments while becoming aware of methods to track and control them. “What’s Lurking in That Building?” features important pedagogical approaches that allow students to learn, which may translate environmental health and basic epidemiological concepts into practical application. Inherent in the design is its ability to encourage critical thinking, authoritative decision-making, and practical problem solving (Science Center 2014). In terms of children learning about the environment, this game captures the content specific and pedagogical approaches identified and emphasized in the literature for teaching and learning (Rudsberg and Öhman 2010).

### Limitations

There are some limitations to this study. Only a small number of children participated from elementary schools in an urban central Adriatic Sea community in Croatia, thus our results may not be generalizable to children in other grades or settings. We surveyed students on their general rather than specific knowledge of epidemiological or environmental health concepts or changes in their knowledge based on the game. Most of the children responding to the survey were from one of the Eco-schools in the region, thus their experience with classes related to environmental health or epidemiology might influence their attitudes about the workshop or game. Additionally, we did not collect data on gender. A recent study showed that elementary school girls scored higher than boys did in a test of action competence from exposure to either an environmental education curriculum or not (Jan Cincera and Krajhanzl 2013). As the number of participants of the workshop was unknown beforehand, we prepared only two complete games. Consequently, the groups of players were larger by two to three students, thus the overall exposure to certain elements of the game may have been reduced for the players involved. We did not collect data from respondents regarding which specific elements of the game they preferred, such as decision-making or problem solving. Additionally, perhaps the children did not have enough of the recommended exposure time to the game to have meaningful learning during the time allotted for game play, which should consist of at least two separate 50-minute class periods (Science Center 2014). Further, the pedagogical content of the game was intended for children from third to sixth grades to help them learn environmental epidemiology concepts. Our pilot study involved children from seventh and eighth grades; nonetheless, these children reported that they learned from playing. Furthermore, the children completed the survey during the workshop, so they may have been compelled to give answers that the organizers would find pleasing. Lastly, although we did not translate any aspects of the game into Croatian, the reading level of the game was at least at a third grade reading level. Even though all of the children had taken mandatory English language classes in elementary school, some of the students may have been at different levels of reading proficiency and comprehension in English.

It is important to note that 85% of the participants were from an Eco-school and that their experiences with environmental health or epidemiology may be greater than those experiences of students from other schools. A game-based curriculum could supplement existing curricula with or without an eco-program by introducing environmental epidemiology for translation into practical application of concepts or development of students into future researchers and teachers.

The “What’s Lurking in that Building?” game appears to be a nascent way to introduce environmental epidemiology to schools with an absence of environmental health or epidemiology in their curricula. Moreover, the problem-solving or decision-making aspects of the game could enrich the learning experience of pupils who already learn these subjects. This study suggests that schools similar to those in this study may benefit from the introduction of this game in that children would enjoy learning about environmental epidemiology. In a future longitudinal study, we could assess the specific concepts of the game (such as problem solving or decision-making) that are linked to behavioral change outcomes. Perhaps behavioral change regarding the environment could extend beyond the classroom to encourage future leaders in sustainable development.

## Data Availability

Availability of data and material: The data that support the findings of this study are openly available in the Open Science Framework.

https://osf.io/fyqa3/.

## Availability of data and material

The data that support the findings of this study are openly available in the Open Science Framework at https://osf.io/fyqa3/.

## Code availability

Not applicable

## Ethics approval

All procedures performed in studies involving human participants were in accordance with the ethical standards of the institutional and/or national research committee (Ethics Committee at the University of Split School of Medicine, reference number 2181-198-03-04-18-0063) and with the 1964 Helsinki declaration and its later amendments or comparable ethical standards.

## Authors’ contributions

Shelly Melissa Pranic conceived and designed the study, collected the data, executed the statistical analyses, and interpreted the results. Tina Batinovic edited and translated the questionnaire and interpreted the results. All authors actively participated in the writing and editing of the manuscript before approving the final draft.

