## Supplementary Materials for "Impact of an environmental epidemiology board game on knowledge, experience, and attitudes among children: A pilot study"

**Author Names:** 1. Shelly Melissa Pranic^1^*, 2. Tina Batinovic^2^

**Affiliations:** ^1^Department of Public Health, University of Split School of Medicine, Soltanska 2, 21000 Split, Croatia

^2^University of Split School of Medicine, Soltanska 2, 21000 Split, Croatia

*Corresponding author: ORCID ID: https://orcid.org/0000-0001-5524-1723

**E-mail Addresses (in order of appearance of the authors from above):**

Supplementary Materials

Supplementary Table 1. Characters and discovery objectives of both the “What's Lurking in Lunch? and “What's Lurking in that Building?” game to help elementary school students discover the source of an illness from an

environmental contaminant.

Supplementary Table 2. Open-ended survey responses from elementary school students regarding a workshop and “What's Lurking in that Building?” game on environmental epidemiology.

Supplementary Questionnaire

ICMJE Disclosure Form A

ICMJE Disclosure Form B

The EQUATOR network STROBE Statement—Checklist of items that should be included in reports of cross-sectional studies

Supplementary Table 1. Characters and discovery objectives of both the “What's Lurking in Lunch? and “What's Lurking in that Building?” game to help elementary school students discover the source of an illness from an environmental contaminant.

| Discovery objectives |  | “What's Lurking in Lunch?”^a^ | | |  | “What's Lurking in that Building?” |
| --- | --- | --- | --- | --- | --- | --- |
| Culprit^b^ |  | Dirty Dan - unhygienic food-handler | | |  | Messy Marko - forgot to dispose of mercury-containing thermometers from an abandoned factory |
|  |  | Filthy Frieda - improperly washes food surfaces and utensils | | |  |  |
|  |  |  |  |  |  | Lame Lina - did not cut tall grass in and around the playground |
|  |  | Contagious Clark - sneezes and coughs without covering his mouth | | |  | Cranky Karlo - forgets to close the dog cages at the pet shop, so people freely pet them |
|  |  | Careless Christine - improperly stores foods | | |  | Careless Kristian - neglected to throw away old chemicals from leaky containers |
|  |  | Shortcut Shelley - fails to cook food fully | | |  |  |
|  |  |  |  |  |  | Sloppy Sime - failed to check well-water contamination at his school |
| Contaminant^c^ |  | *Campylobacter jejuni* | | |  | Arsenic |
|  |  | *Listeria monocytogenes* | | |  | Dog's dandruff |
|  |  | *Norovirus* | |  |  | Ragweed pollen |
|  |  | *Salmonella* | |  |  | Grass pollen |
|  |  | *Staphylococcus aureus* | | |  | Mercury |
|  |  |  |  |  |  | Carbon tetrachloride  *Stachybotrys Chartarum* |

Supplementary Table 1, *continued*

| Location^d^ |  | Hamburger Hamlet | |  |  | Happy Home |
| --- | --- | --- | --- | --- | --- | --- |
|  |  | Burrito Barn | |  |  | Merry Mall |
|  |  | Ice Cream Igloo | |  |  | Fun Factory |
|  |  | Sub Shack | |  |  | Playful Playground |
|  |  | Chicken Coop | |  |  | Classy Classroom |
|  |  | Cafeteria |  |  |  |  |
| Environmental source^e^ |  | Condiments | |  |  | Plants and flowers |
|  |  | Dairy |  |  |  | Animals and insects |
|  |  | Deli meats | |  |  | Indoor items |
|  |  | Pre-cooked/packaged foods | | |  | Cleaning supplies and water |
|  |  | Uncooked produce | |  |  | Home decorations |
|  |  | Undercooked meats | | |  |  |
| Laboratory test^f^ |  | Antibody test | |  |  | Liquid chromatography – mass spectrometry based multi-mycotoxin methods, Quantitative Fluorescent Enzyme Immunoassay |
|  |  | Molecular test | |  |  |  |
|  |  | Selective culture media | | |  |  |
|  |  | Staining |  |  |  |  |
|  |  | Microscopy | |  |  | Immunoglobulin E (IgE) blood test |
|  |  |  |  |  |  | Arsenic test strip |
|  |  |  |  |  |  | Skin prick test |
|  |  |  |  |  |  | Field portable mercury vapor analyzer |

^a^Science Center 2014.

^b^Character responsible for causing illness through improper cleaning or handling of hazardous substances.

^c^Substance responsible for causing illness in individuals; referred to as “pathogen” in “What's Lurking in Lunch?”

^d^Building or surroundings where illnesses occurred.

^e^Source of contaminant; referred to as “food” in “What's Lurking in Lunch?”

^f^Test performed or instrument used to determine the cause of an illness.

Supplementary Table 2. Open-ended survey responses from elementary school students regarding a workshop and “What's Lurking in that Building?”^a^ game on environmental epidemiology.

| **Questions** | |  |  | **Comments about the game and workshop** |
| --- | --- | --- | --- | --- |
| How did you feel about the brief talk on environmental health? | | | |  |
| Why? |  |  |  | It is interesting; we can learn both subjects about taking care of the environment; what I learn, I can apply at home or outside; it's important for many things. |
| Can you give an example? | | |  | We can clean the dust around the house; so people can be healthy. |
| What kind of activities did you enjoy about the “Green is the new black” workshop? | | | |  |
|  |  |  |  | The colors; the game; we built houses; the houses. |

^a^Modified from the original “What's Lurking in Lunch?” game [1].

Supplementary Questionnaire

University of Split

Department of Public Health

“Green is the new black” workshop

Welcome, students! This survey seeks to gather your comments and suggestions about the “Green is the new black” workshop that you have attended today. By completing this short survey (about 10 minutes), will ask questions concerning your experience and attitudes concerning environmental health science and epidemiology. The answers that you provide will be used to encourage the introduction of environmental health science and epidemiology courses into the curriculum of your elementary or high school and others.

Your participation is voluntary.

Please tell us a little about yourself:

What is the name of your school:_________________________________

Which grade are you in for the 2015/2016 school year: ___________

Age:_______

1. How would you describe your classes on environmental issues before the “Green is the new black” workshop?

1 = never had a course in environmental health at school

2 = had some courses on environmental health science at school

3 = environmental health science topics were taught in a biology course at school with other science courses at school

4 = environmental health science topics were taught separately from a biology course at school

2. Did you enjoy this part of the workshop when we discussed environmental issues?

1= did not like

2 = so-so or neutral

3 = liked

4 = loved

3.	How would you describe your epidemiology (tracking the occurrence and control of factors that affect your health) classes before the “Green is the new black” workshop?

1 = never had a course in epidemiology at school

2= had some courses on epidemiology at school

3 = epidemiology topics were taught in a biology course at school or with other science courses at school

4 = epidemiology topics were taught separately from a biology course at school

Supplementary Questionnaire, *continued*

4.	How did you feel about the brief talk about epidemiology?

1 = did not like

2 = so-so or neutral

3 = liked

4 = loved

Why?

__________________________

Can you give an example?

_________________________________________________________________________

5. How much did you learn about environmental health science from the brief talk in the workshop?

1 = nothing

2 = not very much

3 = some useful things

4 = a lot

6. **Please skip to question # 7** if you did not play the “What’s Lurking in That Building?” game.

How much did you learn about environmental health science from the “What’s Lurking in That Building” Game?

1 = nothing

2 = not very much

3 = some useful things

4 = a lot

7. **Please skip to question # 8** if you did not participate in the “Constructing a green building” activity.

How much did you learn about environmental health science from the “Constructing a green building” activity?

1 = nothing

2 = not very much

3 = some useful things

4 = a lot

Supplementary Questionnaire, *continued*

Please write your thoughts about the “Green is the new black” workshop:

8. What kind of activities did you enjoy about the “Green is the new black” workshop?

______________________________________________________________________________________________________________________________________________________

9. Is there anything else you would like to share about the “Green is the new black” workshop?

______________________________________________________________________________________________________________________________________________________

Thank you again for your time!

**ICMJE DISCLOSURE FORM A**

**Date: May 9, 2021**

**Your Name: Shelly Melissa Pranic**

**Manuscript Title: Impact of an environmental epidemiology board game on knowledge, experience, and attitudes among children:**

**A pilot study**

**Manuscript number (if known):__________________________________________________________________**

**In the interest of transparency, we ask you to disclose all relationships/activities/interests listed below that are**

**related to the content of your manuscript. “Related” means any relation with for-profit or not-for-profit third**

**parties whose interests may be affected by the content of the manuscript. Disclosure represents a commitment**

**to transparency and does not necessarily indicate a bias. If you are in doubt about whether to list a relationship/activity/interest, it is preferable that you do so.**

**The following questions apply to the** **author’s relationships/activities/interests as they relate to the current**

**manuscript only.**

**The author’s relationships/activities/interests should be defined broadly. For example, if your manuscript pertains**

**to the epidemiology of hypertension, you should declare all relationships with manufacturers of antihypertensive medication, even if that medication is not mentioned in the manuscript.**

**In item #1 below, report all support for the work reported in this manuscript without time limit. For all other items,**

**the time frame for disclosure is the past 36 months.**

|  |  | **Name all entities with whom you have this relationship or indicate none (add rows as needed)** | | **Specifications/Comments**  **(e.g., if payments were made to you or to your institution)** |
| --- | --- | --- | --- | --- |
| **Time frame: Since the initial planning of the work** | | | | |
| 1 | All support for the present manuscript (e.g., funding, provision of study materials, medical writing, article processing charges, etc.)  **No time limit for this item.** | University of Split, Split, Croatia | | A one-time payment was made to me. |
| **Time frame: past 36 months** | | | | |
| 2 | Grants or contracts from any entity (if not indicated in item #1 above). | X None |  | |
| 3 | Royalties or licenses | X None |  | |
| 4 | Consulting fees | X None |  | |
| 5 | Payment or honoraria for lectures, presentations, speakers bureaus, manuscript writing or educational events | X None |  | |
| 6 | Payment for expert testimony | X None |  | |
| 7 | Support for attending meetings and/or travel | X None |  | |
| 8 | Patents planned, issued or pending | X None |  | |
| 9 | Participation on a Data  Safety Monitoring Board or Advisory Board | X None |  | |
| 10 | Leadership or fiduciary role in other board, society, committee or advocacy group, paid or unpaid | X None |  | |
| 11 | Stock or stock options | X None |  | |
| 12 | Receipt of equipment, materials, drugs, medical writing, gifts or other services | X None |  | |
| 13 | Other financial or non-financial interests | X None |  | |

**Please place an “X” next to the following statement to indicate your agreement:**

**X	I certify that I have answered every question and have not altered the wording of any of the questions on this**

**form.**

**ICMJE DISCLOSURE FORM B**

**Date: May 09, 2021**

**Your Name: Tina Batinovic**

**Manuscript Title: Impact of an environmental epidemiology board game on knowledge, experience, and attitudes among children:**

**A pilot study**

**Manuscript number (if known):__________________________________________________________________**

**In the interest of transparency, we ask you to disclose all relationships/activities/interests listed below that are**

**related to the content of your manuscript. “Related” means any relation with for-profit or not-for-profit third**

**parties whose interests may be affected by the content of the manuscript. Disclosure represents a commitment**

**to transparency and does not necessarily indicate a bias. If you are in doubt about whether to list a relationship/activity/interest, it is preferable that you do so.**

**The following questions apply to the author’s relationships/activities/interests as they relate to the current**

**manuscript only.**

**The author’s relationships/activities/interests should be defined broadly. For example, if your manuscript pertains**

**to the epidemiology of hypertension, you should declare all relationships with manufacturers of antihypertensive medication, even if that medication is not mentioned in the manuscript.**

**In item #1 below, report all support for the work reported in this manuscript without time limit. For all other items,**

**the time frame for disclosure is the past 36 months.**

|  |  | **Name all entities with whom you have this relationship or indicate none (add rows as needed)** | | **Specifications/Comments**  **(e.g., if payments were made to you or to your institution)** |
| --- | --- | --- | --- | --- |
| **Time frame: Since the initial planning of the work** | | | | |
| 1 | All support for the present manuscript (e.g., funding, provision of study materials, medical writing, article processing charges, etc.)  **No time limit for this item.** | University of Split, Split, Croatia | | A one-time payment was made to Shelly Melissa Pranic. |
| **Time frame: past 36 months** | | | | |
| 2 | Grants or contracts from any entity (if not indicated in item #1 above). | X None |  | |
| 3 | Royalties or licenses | X None |  | |
| 4 | Consulting fees | X None |  | |
| 5 | Payment or honoraria for lectures, presentations, speakers bureaus, manuscript writing or educational events | X None |  | |
| 6 | Payment for expert testimony | X None |  | |
| 7 | Support for attending meetings and/or travel | X None |  | |
| 8 | Patents planned, issued or pending | X None |  | |
| 9 | Participation on a Data  Safety Monitoring Board or Advisory Board | X None |  | |
| 10 | Leadership or fiduciary role in other board, society, committee or advocacy group, paid or unpaid | X None |  | |
| 11 | Stock or stock options | X None |  | |
| 12 | Receipt of equipment, materials, drugs, medical writing, gifts or other services | X None |  | |
| 13 | Other financial or non-financial interests | X None |  | |

**Please place an “X” next to the following statement to indicate your agreement:**

**X	I certify that I have answered every question and have not altered the wording of any of the questions on this**

**form.**

STROBE Statement—Checklist of items that should be included in reports of ***cross-sectional studies***

|  | Item No | Recommendation | Page No |
| --- | --- | --- | --- |
| **Title and abstract** | 1 | (*a*) Indicate the study’s design with a commonly used term in the title or the abstract | 1 |
|  |  | (*b*) Provide in the abstract an informative and balanced summary of what was done and what was found | 1 |
| Introduction | | | |
| Background/rationale | 2 | Explain the scientific background and rationale for the investigation being reported | 2-4 |
| Objectives | 3 | State specific objectives, including any prespecified hypotheses | 4 |
| Methods | | | |
| Study design | 4 | Present key elements of study design early in the paper | 5 |
| Setting | 5 | Describe the setting, locations, and relevant dates, including periods of recruitment, exposure, follow-up, and data collection | 5-6 |
| Participants | 6 | (*a*) Give the eligibility criteria, and the sources and methods of selection of participants | 5-6 |
| Variables | 7 | Clearly define all outcomes, exposures, predictors, potential confounders, and effect modifiers. Give diagnostic criteria, if applicable | 5-6 |
| Data sources/ measurement | 8* | For each variable of interest, give sources of data and details of methods of assessment (measurement). Describe comparability of assessment methods if there is more than one group | 5-6 |
| Bias | 9 | Describe any efforts to address potential sources of bias | 6 |
| Study size | 10 | Explain how the study size was arrived at | N/A |
| Quantitative variables | 11 | Explain how quantitative variables were handled in the analyses. If applicable, describe which groupings were chosen and why | 7 |
| Statistical methods | 12 | (*a*) Describe all statistical methods, including those used to control for confounding | 7 |
|  |  | (*b*) Describe any methods used to examine subgroups and interactions | N/A |
|  |  | (*c*) Explain how missing data were addressed | N/A |
|  |  | (*d*) If applicable, describe analytical methods taking account of sampling strategy | 7 |
|  |  | (*e*) Describe any sensitivity analyses | N/A |
| Results | | | |
| Participants | 13* | (a) Report numbers of individuals at each stage of study—eg numbers potentially eligible, examined for eligibility, confirmed eligible, included in the study, completing follow-up, and analysed | 8 |
|  |  | (b) Give reasons for non-participation at each stage | N/A |
|  |  | (c) Consider use of a flow diagram | N/A |
| Descriptive data | 14* | (a) Give characteristics of study participants (eg demographic, clinical, social) and information on exposures and potential confounders | 8 |
|  |  | (b) Indicate number of participants with missing data for each variable of interest | N/A |
| Outcome data | 15* | Report numbers of outcome events or summary measures | 8-11 |
| Main results | 16 | (*a*) Give unadjusted estimates and, if applicable, confounder-adjusted estimates and their precision (eg, 95% confidence interval). Make clear which confounders were adjusted for and why they were included | N/A |
|  |  | (*b*) Report category boundaries when continuous variables were categorized | N/A |
|  |  | (*c*) If relevant, consider translating estimates of relative risk into absolute risk for a meaningful time period | N/A |
| Other analyses | 17 | Report other analyses done—eg analyses of subgroups and interactions, and sensitivity analyses | N/A |
| Discussion | | | |
| Key results | 18 | Summarise key results with reference to study objectives | 12-13 |
| Limitations | 19 | Discuss limitations of the study, taking into account sources of potential bias or imprecision. Discuss both direction and magnitude of any potential bias | 13-14 |
| Interpretation | 20 | Give a cautious overall interpretation of results considering objectives, limitations, multiplicity of analyses, results from similar studies, and other relevant evidence | 14 |
| Generalisability | 21 | Discuss the generalisability (external validity) of the study results | 14-15 |
| Other information | | | |
| Funding | 22 | Give the source of funding and the role of the funders for the present study and, if applicable, for the original study on which the present article is based | 1, 15 |

*Give information separately for exposed and unexposed groups.
